# Estimating the serotype-specific association between the concentration of vaccine-induced serum antibodies and protection against pneumococcal colonization

**DOI:** 10.1101/2024.10.17.24315707

**Authors:** Anabelle Wong, Joshua L. Warren, Laura Fitch, Stephanie Perniciaro, Ron Dagan, Daniel M. Weinberger

## Abstract

**Background:** Pneumococcal conjugate vaccines (PCVs) offer indirect protection by reducing pneumococcal colonization in the vaccinated children and thus transmission. As higher-valency PCVs may trigger a weaker immune response, it is important to understand how differences in immunogenicity between PCVs translate to effectiveness against colonization.

**Methods:** We estimated the serotype-specific relationship between the concentration of vaccine-induced serum immunoglobulin G (IgG) and protection against colonization using a hierarchical Bayesian model with the longitudinal data from a randomized controlled trial in Israel. Then, we combined these estimates with the summary-level immunogenicity data (geometric mean concentration and 95% confidence intervals) from head-to-head clinical trials comparing PCV13 vs. PCV7, PCV 15 vs. PCV13, and PCV20 vs. PCV13 to infer the relative effectiveness of higher-valency PCVs against colonization.

**Results:** The hierarchical Bayesian model predicted that the risk of colonization increased as serum IgG decreased, and the association differed by serotype. Our approach estimated higher-valency PCVs to have lower vaccine effectiveness against colonization with some serotypes: 14 and 23F across comparisons; 4 when comparing PCV13 with PCV7 and comparing PCV20 with PCV13; 5, 6A, 6B 7F, 19A, and 19F when comparing PCV15 and PCV20 with PCV13, and additionally 1, 9V and 18C when comparing PCV20 with PCV13.

**Conclusions:** These findings suggest that while new PCVs might provide sufficient protection against severe disease, protection against transmission might be somewhat reduced for some serotypes. The overall impact should be evaluated in the local context and further monitoring is critical to evaluate the impact of these changes in the coming years.

## Introduction

The introduction of pneumococcal conjugate vaccines (PCVs) has substantially reduced the burden of invasive pneumococcal diseases (IPD) in young children [1]. PCVs offer direct protection to vaccinated children and indirect protection to unvaccinated children and older adults by reducing pneumococcal colonization in children and, thus, the transmission from children to their contacts in daily life [2,3]. This indirect protection also enhances the protection of vaccinated children.

PCVs trigger the production of anticapsular immunoglobulin G (IgG). The PCV-induced concentration of these antibodies in serum is used as a correlate of protection against IPD. Immunogenicity measured in randomized controlled trials (RCTs) provides a basis of comparison between new PCVs and established PCVs and has been used as the basis for licensing new PCVs. Based on a meta-analysis [4], a serum IgG concentration of 0.35 μg/mL was set as the threshold of protection against IPD, mainly for the purpose of licensing new PCVs when efficacy studies are not feasible [5]. In reality, the threshold required for protection differs by serotype for IPD [6]. Protection against acquisition of nasopharyngeal colonization scales with higher antibody concentrations rather than having a fixed threshold associated with protection [7].

As new higher-valency PCVs become available, a reduction in the strength of the immune response may weaken protection against the acquisition of colonization for some serotypes. As PCV valency increased, the antibody levels for some serotypes were somewhat lower [8]. While the antibody concentration of the new PCVs is likely sufficient to protect against IPD [9], effectiveness against colonization may be eroded, as a considerably stronger immune response is required to protect against colonization than diseases [7,10].

There is an acute need to understand how differences in immunogenicity between PCVs translate to effectiveness against colonization. Previous work has mostly focused on assessing the association between post-primary antibody responses and risk of colonization in the first year of life [7]. However, transmission of pneumococcus is thought to be driven by preschool and school-aged children [11,12]. Therefore, estimating the relationship between immunogenicity and risk of colonization in older children after receipt of the booster dose is particularly important for understanding the differences in immunogenicity between PCVs for transmission. There have been attempts to assess the relationship between post-booster antibody responses and risk of colonization by serotype and risk group; however, the estimates were unstable for many serotypes due to scarce events [10]. Re-analyzing these data using advanced analytical methods, such as hierarchical Bayesian statistical models, can help stabilize the estimated serotype-specific relationship between serum antibody levels and protection against colonization [13].

In this study, we first estimated the relationship between the concentration of serum IgG and risk of colonization by serotype by re-analyzing data from a RCT in Israel using a hierarchical Bayesian model. Then, we combined these estimates with summary-level immunogenicity data from head-to-head clinical trials to infer the relative effectiveness of new PCVs against colonization (Figure 1).

**Figure 1.**
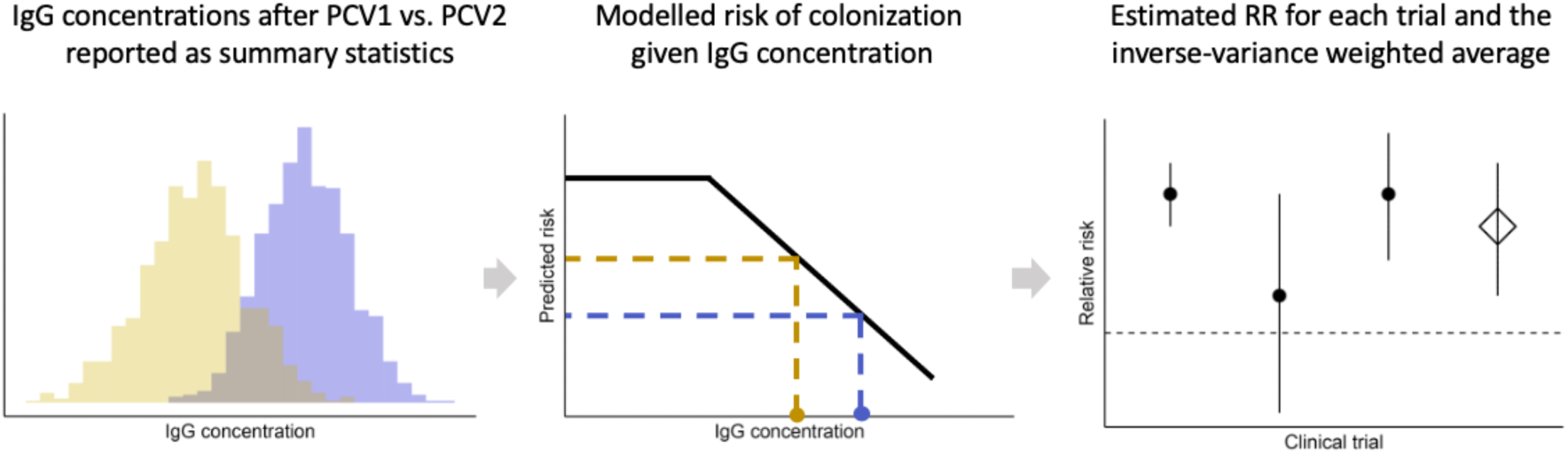
A schematic showing the modeling workflow. The serotype-specific immunogenicity data from head-to-head clinical trials comparing a higher-valency PCV with an older, lower-valency PCV (left panel), summarized and reported as mean and 95% confidence intervals, are publicly available. Combining these data with a hierarchical Bayesian model fitted to the longitudinal data on post-PCV immunogenicity and colonization acquisition from one randomized controlled trial [10,14] (middle panel) gives the relative risk of colonization for each head-to-head trial. The estimated relative risk of colonization for all included head-to-head trials for a specific comparison (e.g., PCV13 vs. PCV7) were pooled by inverse-variance weighting (right panel).

## Methods

### Data

Two sets of data were used in this study: (1) longitudinal data on post-PCV immunogenicity and colonization from a RCT, and (2) summary-level immunogenicity data from head-to-head clinical trials.

The first data set was described in [10,14]. The serotype-specific IgG concentration was measured using standardized enzyme-linked immunosorbent assay (ELISA) 1 month after the children received their toddler booster dose (i.e., at age 13 months) with either PCV7 or PCV13. Ethnicity of the children (Bedouin, Jewish) was recorded. Nasopharyngeal swabs were collected and cultured at 2, 4, 6, 7, 12, 13, 18, and 24 months of age to detect acquisition of colonization. Positive samples were serotyped by Quellung reaction. Pneumococcal acquisition was recorded as a binary outcome for each of the thirteen serotypes included in PCV13 (4, 6B, 9V, 14, 18C, 19F, 23F, 1, 3, 5, 6A, 7F, 19A). For post-booster IgG at age 13 months, we examined colonization prior to age 13 months to exclude baseline colonization, and if a participant was found to be colonized by a given PCV13 serotype, the subsequent observations for that same serotype were censored (eAppendix 1). In addition to the post-booster IgG data, we also analyzed the association between serotype-specific IgG concentration 1 month after the primary series (i.e., at age 7 months) and risk of colonization after the booster dose and presented the results in eFigure 10.

The second set of data came from head-to-head clinical trials comparing PCV13 vs. PCV7, PCV15 vs. PCV13, and PCV20 vs. PCV13. A scoping review was conducted to provide a list of studies evaluating the immunogenicity of PCVs [15]. Head-to-head clinical trials with results on the post-primary or post-booster IgG geometric mean concentration (GMC) for the vaccines being compared were selected (see eFigures 2–4 for the flowcharts of study inclusion). For the selected studies, the summary-level immunogenicity data (GMCs and 95% confidence intervals [CIs]) were extracted from WISSPAR.com and ClinicalTrials.gov.

**Figure 2.**
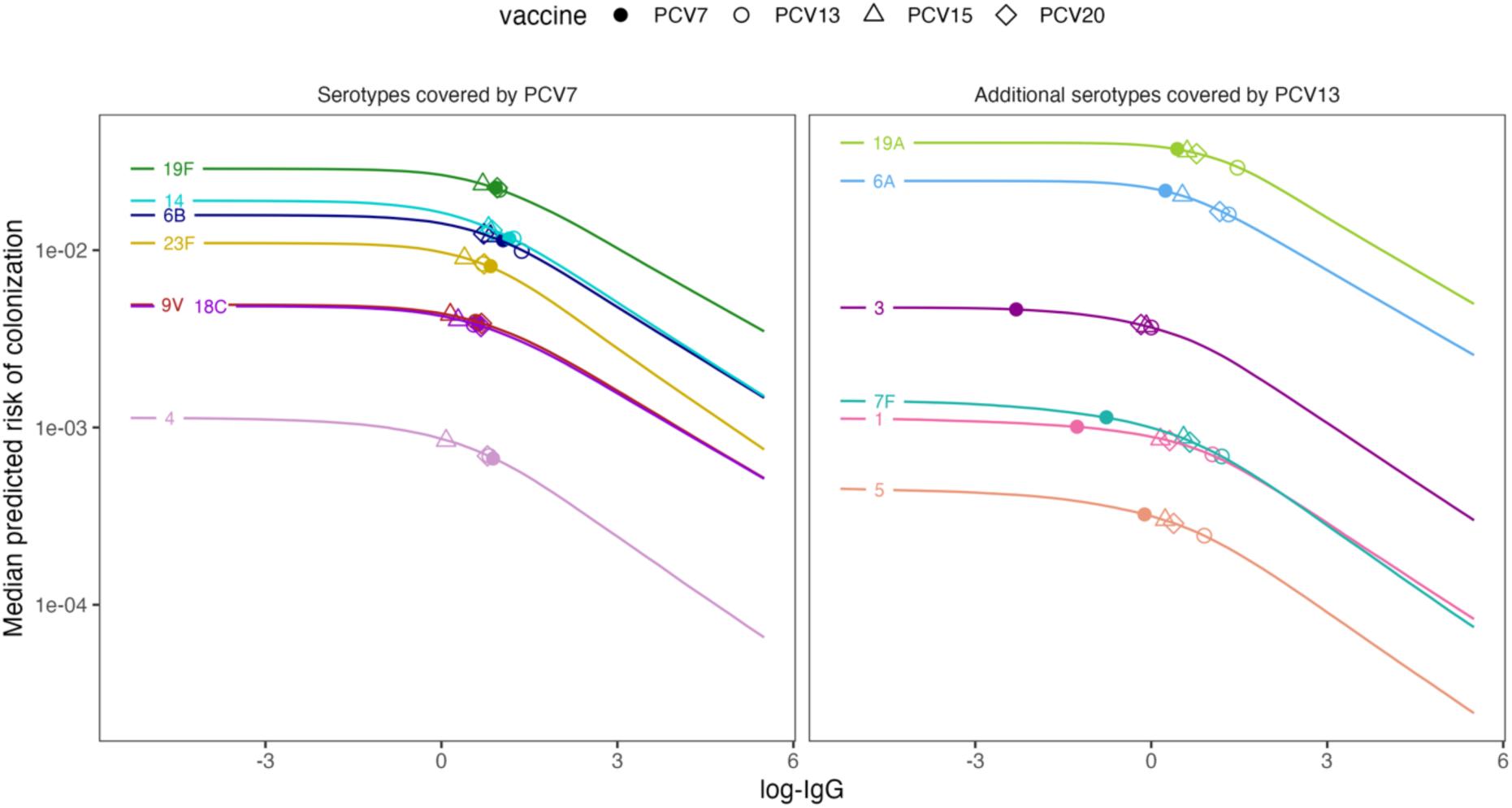
The predicted probability of colonization by antibody concentration for each serotype. The median predicted risk of colonization from the hierarchical Bayesian model for each serotype (left: serotypes covered by PCV7 – 4, 6B, 9V, 14, 18C, 19F, 23F; right: additional serotypes covered by PCV13 – 1, 3, 5, 6A, 7F, 19A) is plotted against the concentration of serum antibodies. The points indicate the antibody concentrations achieved by the booster dose of PCV7 (filled circle), PCV13 (open circle), PCV15 (open triangle), and PCV20 (open diamond) as an inverse-variance weighted average of the immunoglobulin G (IgG) geometric mean concentration (GMC) reported by the head-to-head clinical trials. The y-axis is displayed in the log10 scale.

### Risk of colonization and serum IgG concentration

Pneumococcal vaccines primarily affect colonization by preventing the acquisition of pneumococcus rather than shortening the duration of colonization. Given the longitudinal data that we have, with measurements at just three time points, we cannot assess acquisition rates. Therefore, we assess the risk of being colonized in each sample as a function of antibody IgG concentration at 13 months of age. To model this, we assumed that for a given serotype, the risk of colonization decreased with increasing serum IgG concentration following a log-log relationship. We also assumed that there was a minimum amount of antibody needed before any reduction in risk is observed. We developed a hierarchical Bayesian change point model to allow for the sharing of information across different serotypes during model fitting, resulting in more robust estimates of serotype-specific relationships.

Specifically, we model the probability of individual *i* being colonized with the serotype *j* such that

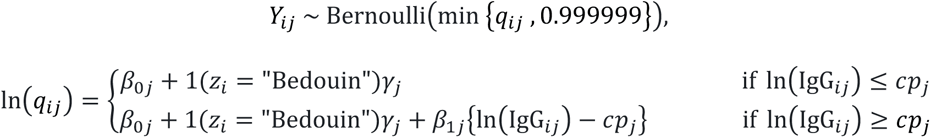

where 𝑌_ij_ is the binary outcome describing if person *i* was colonized with serotype *j* or not; 𝑞_ij_ is the probability of colonization with serotype *j* for person *i*; 𝛽_0j_ is the serotype-specific intercept parameter; 𝛽_1j_ is the serotype-specific slope parameter describing the association between ln(IgG_ij_) and ln(𝑞_ij_); 𝑐𝑝_j_ is the serotype-specific change point parameter; and 𝛾_j_ is the serotype-specific intercept parameter for the Bedouin ethnicity where 𝑧_i_ represents the ethnicity category for person *i* and 1(.) is an indicator function that returns a value of one when the input statement is true and a value of zero when it is false. We include the 0.999999 correction factor to ensure that the modeled probabilities do not exceed 1, a possibility when using the log link function for 𝑞_ij_ without further constraints.

To encourage sharing of information across serotypes during estimation, we assign the following hierarchical prior distributions to the intercepts, slopes, and change points:

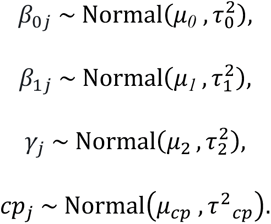

To complete the model specification, we specified prior distributions for the hyperparameters. Specifically, 𝜇*_0_*, 𝜇_1_ and 𝜇_2_ are normally distributed with mean 0 with variance 10^4^; 𝜇_cp_ is normally distributed with mean equal to the log-IgG and variance equal to the variance of log-IgG from the observed range of post-PCV serum IgG concentration [10,14]; and 𝜏_0_^2^, 𝜏_1_^2^, 𝜏_2_^2^ and 𝜏_cp_^2^ have independent inverse gamma (0.01, 0.01) distributions.

The model was fitted to the longitudinal data on post-PCV immunogenicity and colonization from [10,14] to estimate the serotype-specific intercepts, change points, slopes, and ethnicity-specific intercepts. Details of the model fitting in the Bayesian setting are given in eAppendix 2. As sensitivity analysis, we tested a less informative prior for 𝜇_cp_ (eAppendix 3). We also tested a simpler linear model (without change point) including random effects for ethnic groups (eAppendix 4). The estimates of the relationship between IgG concentration and risk of colonization for the Bedouin and Jewish groups were not notably different in preliminary analyses (eFigure 10); therefore, we only accounted for the baseline colonization risk difference by including the ethnicity-specific intercepts. This assumes that the relative reductions in risk of colonization achieved with changes in antibody concentration were the same between the ethnic groups, but the baseline risk of colonization differed.

### Relative vaccine efficacy

We identified head-to-head clinical trials comparing PCV13 vs. PCV7, PCV 15 vs. PCV13, and PCV20 vs. PCV13 from a previously conducted scoping review [15] and extracted the summary-level data for the GMC measured using ELISA at 1 month post-booster. For trials involving PCV15, only GMC measured by electrochemiluminescence (ECL) was reported. Therefore, we applied the conversion from ECL-measured GMC to ELISA-measured GMC using [16,17] (eAppendix 5) to ensure comparability of the GMC.

Using the fitted hierarchical Bayesian model, we predicted the risk of colonization with a given serotype for each reported GMC. For the head-to-head comparison in each trial, we divided the predicted risk from a higher-valency PCV by the predicted risk of a lower-valency PCV to obtain the relative risk. Finally, we pooled the estimated relative risk across trials using inverse-variance weights (eAppendix 6). The uncertainty of the estimates came from the posterior samples of the hierarchical Bayesian model and the reported variance of the GMC. With the relative risk of colonization, we can obtain the relative vaccine efficacy (VE) calculated as 1 – RR = VE. In other words, the higher the RR, the lower the relative VE. This can also be interpreted in terms of absolute vaccine effectiveness. For example, assuming that PCV7 confers 60% (RR of 0.4) protection against colonization by a serotype [18,19], an estimated RR of 1.10 when comparing PCV13 with PCV7 can be interpreted as 10% reduction in the absolute VE for the higher-valency PCV, meaning, PCV13 has an absolute VE of 56% (1 – 0.4*1.1). If the RR for PCV20 vs PCV13 was 1.2, we would extend this by calculating the overall VE=47% (1 – 0.4*1.1*1.2).

## Results

### Overview of data

After excluding baseline colonization episodes and censoring colonization episodes of the same serotype following the first event, there were 28192 measurements (Bedouin 10033; Jewish 18159) from 2345 samples, drawn from 896 individuals (Bedouin 323; Jewish 573) in the longitudinal data on post-PCV immunogenicity and colonization [10,14] (eTable 1). Among the included observations, there were 220 newly acquired colonizations (Bedouin 98; Jewish 122).

After identifying registered head-to-head clinical trials for PCV13v7 (n=19), PCV15v13 (n=19), and PCV20v13 (n=13), we extracted the GMCs with 95% CIs from completed studies for post-primary (PCV13v7: n=5; PCV15v13: n=8; PCV20v13: n=3) and post-booster (PCV13v7: n=5; PCV15v13: n=10; PCV20v13: n=4) IgG against thirteen PCV13 serotypes (eFigures 2–4).

### The relationship between risk of colonization and antibody concentration

When fitted to the longitudinal data on post-PCV immunogenicity and colonization from a RCT in Israel, the model estimated a decline in risk of colonization with increasing serum IgG concentration. For most serotypes (6B, 9V, 14, 18C, 19F, 23F, 3, 6A, 19A), the decline in risk only became more apparent above certain serum IgG concentration, suggesting a potential minimal concentration necessary for protection against colonization; for other serotypes (4, 1, 5, 7F), the predicted risk of colonization declined more consistently with increasing serum IgG concentration across the range of observed values (eFigure 5–6, Figure 2). Notably, the latter serotypes had few colonization data points (eTable 1). Although the change points appeared slightly more subtle when using a less informative prior for the hyperparameter 𝜇_cp_ in the sensitivity analysis (eFigure 7), the overall shape of the risk curves and the uncertainty remained similar (eFigure 8). The baseline risk of colonization was higher in the Bedouin group than in the Jewish groups (eFigure 5) and differed by serotype (Figure 2).

The linear model showed no evidence for differential relative reductions in risk of colonization between the ethnic groups (eFigure 10). This model predicted a higher-valency PCV to be associated with a higher risk of colonization with most serotypes for both Bedouin and Jewish children considering the post-booster but not the post-primary IgG concentration (eFigure 10), suggesting such effects may only become apparent after the booster dose. In general, the estimated relative risks of colonization from the change point model were similar to that from the linear model, but with larger uncertainty intervals (eFigure 11). The inference of relative risk of colonization focuses on the post-booster results from the change point model.

### Relative risk of colonization with the 7 shared serotypes

Across the three head-to-head comparisons, the risk of colonization with serotype 14 was 7–8 percent higher when using a higher-valency PCV compared with using a lower-valency PCV (Figure 3; PCV13v7: RR=1.07, 95% credible intervals 1.01–1.13; PCV15v13: 1.08, 1.04–1.12; PCV20v13: 1.07, 1.02–1.13). If vaccine effectiveness against colonization for PCV7 was 60%, this reduction in immunogenicity would result in a VE against colonization of 57% for PCV13 and 54% for PCV15 and for PCV20 (eFigure 15). Another serotype that posed a higher risk of colonization when using a higher-valency PCV was serotype 23F, where the risk was 4–26 percent higher compared with using a lower-valency PCV (PCV13v7: 1.09, 1.01–1.18; PCV15v13: 1.04, 1.00–1.08; PCV20v13: 1.26, 1.11–1.43), with a high degree of uncertainty for PCV13 vs PCV7 and PCV20 vs PCV13. This would represent a VE against colonization of 56% for PCV13, 55% for PCV15, and 45% for PCV20, assuming PCV7 has 60% vaccine effectiveness against colonization with serotype 23F (eFigure 15).

**Figure 3.**
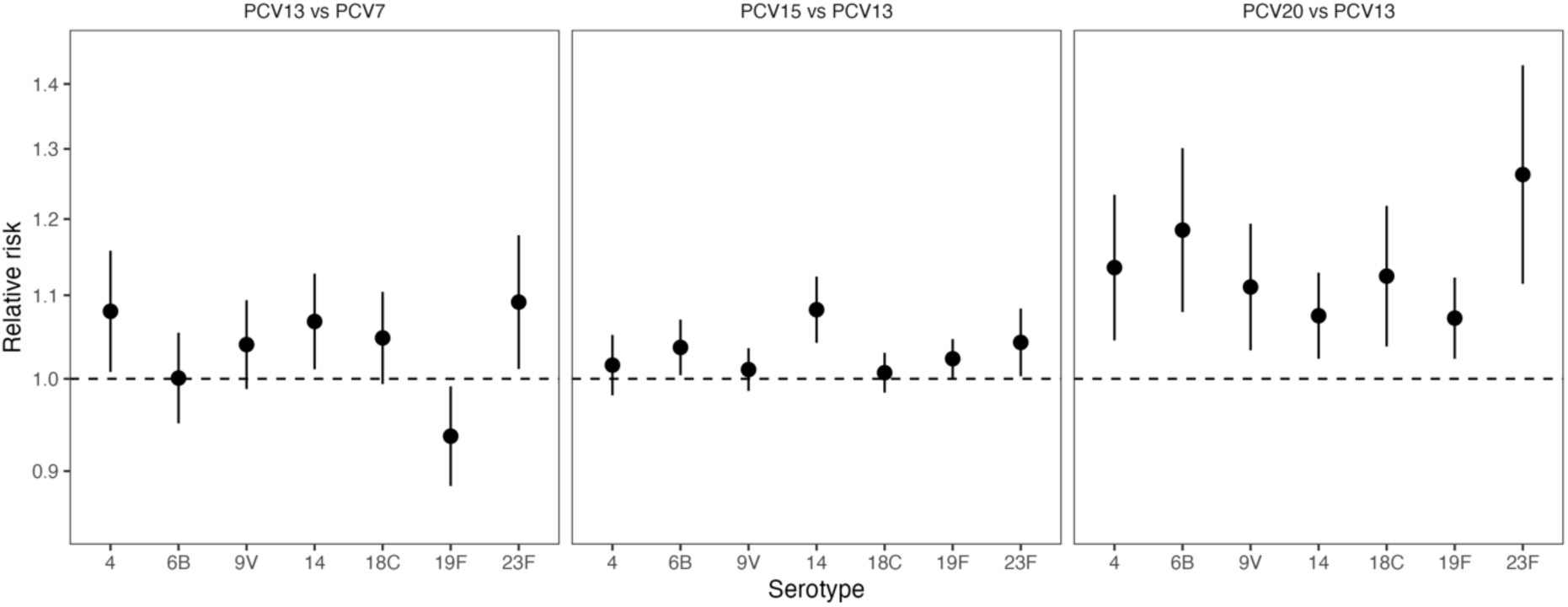
Relative risk of colonization with the 7 shared serotypes for PCV13 vs. PCV7, PCV15 vs. PCV13, and PCV20 vs. PCV13. The estimated relative risk of colonization with the seven serotypes commonly covered by PCV7, PCV13, PCV15, and PCV20 (4, 6B, 9V, 14, 18C, 19F, 23F), when comparing PCV13 with PCV7 (left panel), PCV15 with PCV13 (middle panel), and PCV20 with PCV15 (right panel) is displayed as mean (point) with 95% credible intervals (error bars). The y-axis is displayed in the log10 scale. A relative risk above 1 (indicted by the dashed line) represents a higher risk of colonization when using the higher-valency PCVs.

**Figure 4.**
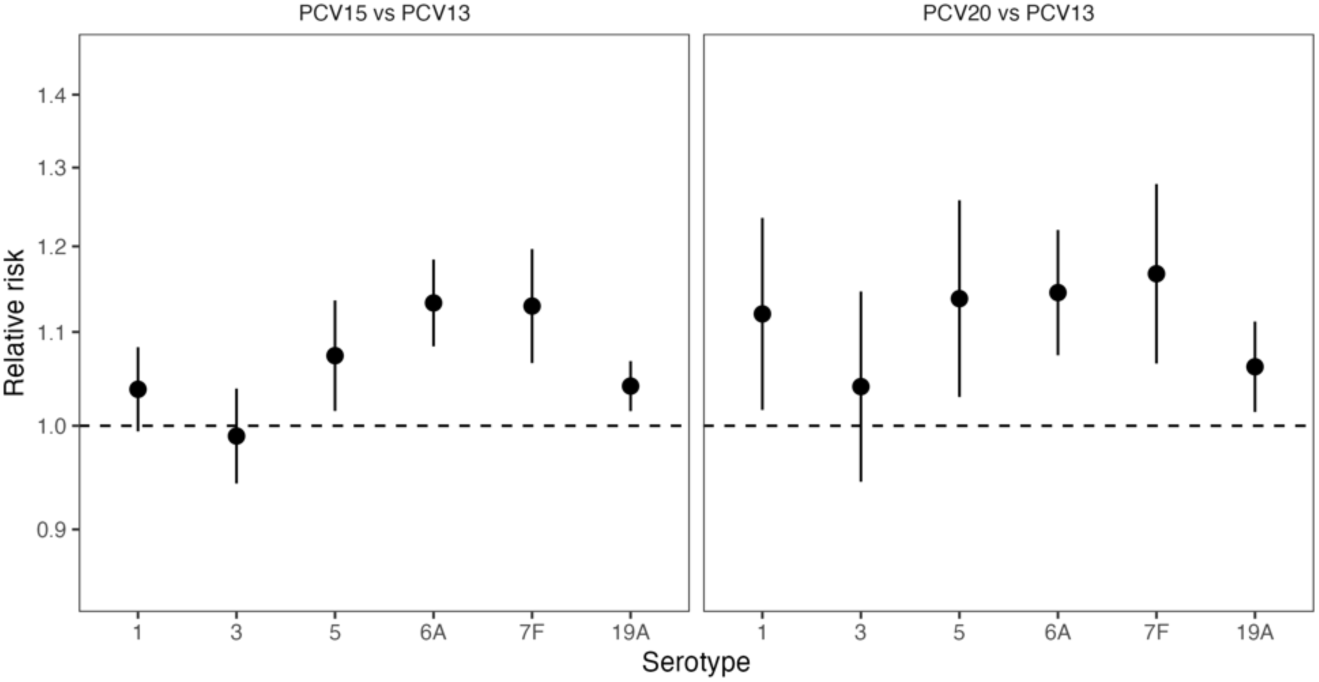
Relative risk of colonization with 6 additional shared serotypes PCV15 vs. PCV13 and PCV20 vs. PCV13. The estimated relative risk of colonization with the six additional serotypes commonly covered by PCV13, PCV15, and PCV20 but not by PCV7 (1, 3, 5, 6A, 7F, 19A), when comparing PCV15 with PCV13 (left panel), and PCV20 with PCV15 (right panel) is displayed as mean (point) with 95% credible intervals (error bars). The y-axis is displayed in the log10 scale. A relative risk above 1 (indicted by the dashed line) represents a higher risk of colonization when using the higher-valency PCVs.

For serotype 6B, the risk of colonization was higher when using PCV15 compared with PCV13 (1.03. 1.00–1.07) and when using PCV20 compared with PCV13 (1.18, 1.08–1.30). Assuming 60% vaccine effectiveness against colonization for PCV7, this would translate to 59% effectiveness for PCV15 and 53% effectiveness for PCV20 (eFigure 15). There was little evidence for a difference in risk for PCV13 vs PCV7 for serotype 6B (1.00, 0.95–1.05). The risk of colonization with serotype 4 was higher when using PCV13 compared with PCV7 (1.08, 1.01–1.16) and when using PCV20 compared with PCV13 (1.14, 1.04–1.23), but not when using PCV15 compared with PCV13 (1.02, 0.98–1.05).

For all of the other serotypes, the differences in risk of colonization for PCV13 vs PCV7 and PCV15 vs PCV13 were not notably different, except for serotype 19F, for which the risk of colonization was lower with PCV13 compared with PCV7 (0.94, 0.99–0.99) and slightly higher with PCV15 compared with PCV13 (1.02, 1.00–1.05) (Figure 3).

Comparing PCV20 and PCV13, the risks of colonization with all serotypes were higher when using the higher-valency PCV, though the uncertainty was higher for some of the estimates (9V: 1.11, 1.03–1.19; 18C: 1.12, 1.03–1.22) than others (19F: 1.07, 1.02–1.12).

### Relative risk of colonization with the 6 additional PCV13 serotypes

For both PCV15 vs PCV13 and PCV20 vs PCV13, the risks of colonization with serotypes 5, 6A, 7F, and 19A were higher when using a higher-valency PCV. Comparing PCV15 and PCV13, the risks of colonization with serotypes 6A and with 7F were both 13 percent higher when using the higher-valency PCV (Figure 4; 6A: 1.13, 1.08–1.18; 7F: 1.13, 1.07–1.20). For serotype 6A, there has been cross-reactivity described with serotype 6B, so these results should be interpreted relative to the VE obtained from cross-reactivity with 6B in PCV7 or PCV13. The higher risks of colonization with these serotypes when using the higher-valency PCV were more pronounced when comparing PCV20 and PCV13, but with higher uncertainty (6A: 1.14, 1.07–1.22; 7F: 1.17, 1.07–1.28). For serotypes 19A and 5, the risks of colonization were estimated to be 4–6 percent and 7–14 percent higher respectively when using the higher-valency PCV in both comparisons, with higher uncertainty for serotype 5 (PCV15v13: 1.07, 1.02–1.14; PCV20v13: 1.14, 1.03–1.26) than serotype 19A (PCV15v13: 1.04, 1.01–1.07; PCV20v13: 1.06, 1.01–1.11). While the risk of colonization with serotype 1 when using a higher-valency PCV was higher in PCV20 vs PCV13 (1.12, 1.02–1.24), the risk of colonization with serotype 3 was not notably different when using a higher-valency PCV for both PCV15 vs PCV13 and PCV20 vs PCV13 in the main analysis. When using a less informative prior for the hyperparameter 𝜇_cp_ in the sensitivity analysis, the protection against colonization with serotype 3 was estimated to be better for PCV15 than PCV13 and worse for PCV20 than PCV13 (eFigure 9).

## Discussion

Using a hierarchical Bayesian model, we re-analyzed longitudinal data on post-PCV immunogenicity and colonization from a RCT [10,14] to estimate the serotype-specific relationship between the concentration of serum IgG and the risk of colonization. These findings suggest that for the PCV15 and PCV20, there might be a modest drop in effectiveness against colonization, which further erodes the effectiveness compared to PCV7. Effectiveness against colonization overall is still moderate, and further monitoring and modeling are needed to determine the implications of these changes in immunogenicity for suppressing colonization of PCV-targeted serotypes. In a population with a well-established PCV program, it is possible that these modest differences in risk of colonization will not have a population-level impact. Nonetheless, these data suggest that there could be tradeoffs between maintaining population-level protection and increasing the number of serotypes covered that need to be quantified and monitored when comparing different PCVs.

Compared to a previous study using these data [14], in which a logistic regression model was fitted for each serotype separately, a hierarchical Bayesian model harnessed information from data across serotypes and has the advantage of stabilizing the estimates for serotypes with sparse count of colonization events [13]. Other recent work has estimated the relationship between immunogenicity and seroincidence of pneumococcus [7]. Those studies have focused on the period after completing the primary series but before the receipt of the booster dose [7,20]. While there is a correlation between post-primary and post-booster responses, there are differences in the responses between these time points, and differences among serotypes are generally less pronounced after receipt of the booster dose. Since controlling carriage in older children is key for maintaining indirect protection, understanding the immune response during this post-booster period is critical.

Our model predicted that the risk of colonization increased as serum IgG decreased, and the association differed by serotype (Figure 2) but not by ethnicity (eFigure 10), as shown in [14] for eight serotypes (PCV7 serotypes plus 6A), and for five additional serotypes (1, 3, 5, 7F, 19A) in our study. Specifically, our model predicted that the risk of colonization decreased readily with increasing serum IgG for serotypes 1, 4, 5, and 7F and less so for serotypes 6B, 9V, 14, 18C, 19F, and 23F (eFigure 6), which echoes the findings in a study using seroincidence data to estimate the serotype-specific correlates of protection against colonization [7]. Furthermore, the results from our model suggested that a higher concentration of serum IgG may be necessary to protect against colonization with serotypes 6B, 14, 19F, and 23F, as observed in [7], and with three additional three serotypes – 3, 6A, and 19A – in our study. These results may help explain the limited indirect effects of PCVs against some serotypes, such as serotype 3, 4, 19A, and 19F in Germany [21], although future studies are required to translate the protection against colonization in children to the changes in risk in other age groups and one should remain cautious about extrapolating the results from one population to another.

Combining the estimated risk curves from our model with the summary-level immunogenicity data from head-to-head clinical trials, we inferred the relative VE comparing new PCVs with established PCVs. This relative VE reflects the protection against colonization in the vaccinated children and is related to VE against mucosal infections like acute otitis media (AOM), given the association between carriage and AOM for certain serotypes [22]. In general, there is a dependence between VE against diseases and VE against colonization as nasopharyngeal colonization is a prerequisite to diseases [23].

Our approach estimated higher-valency PCVs to have lower VE than lower-valency PCVs against colonization with some serotypes. The lower VE by higher-valency PCVs was observed for serotypes 4, 14, 5, 6A, and 7F when comparing PCV15 with PCV13, and PCV20 with PCV13. In addition, PCV20 was estimated to have lower VE also against colonization with serotypes 6B and 23F. Therefore, it will be essential to remain vigilant in monitoring serotype-specific carriage and disease when switching to higher-valency PCVs.

There are several limitations in our study. First, we base our model on serum IgG, which provides a correlate of protection but does not reflect all mechanisms of protection, including T cell response [24], neutrophil killing [25], and mucosal immunity [26,27], which are difficult to measure. Second, the longitudinal data used for fitting the hierarchical Bayesian model come from one location, which limits the generalizability of our model to other populations; nevertheless, estimates for two ethnic groups were compared using the linear model and we saw no evidence for differential protection against colonization by ethnicity after adjusting for differences in baseline risk of colonization (eFigure 10). Third, our model relies on data from RCTs, which represent scenarios of healthy infants with high immunization schedule compliance and thus cannot be generalized to the whole population. It would be helpful to validate our model against real-world data should they become available as the newly approved higher-valency PCVs become more widely used. Finally, although a hierarchical model can reduce noise and stabilize the estimates by sharing information across serotypes, it may introduce bias for serotypes with behaviors markedly different from others, for example, serotype 3. A hybrid model that incorporates a hierarchical model for similar serotypes with the flexibility to model unique serotypes independently may help address this limitation.

Using a novel approach to compare higher-valency PCVs with established ones, our study offers directly interpretable estimates of the relative risk of colonization using different PCVs. According to our estimates, higher-valency PCVs tended to have lower VE against colonization with certain serotypes, for example, serotype 14 and 23F across comparisons; serotype 4 when comparing PCV13 with PCV7 and comparing PCV20 with PCV13; serotypes 5, 6A, 6B 7F, 19A, and 19F when comparing PCV15 and PCV20 with PCV13, and additionally serotypes 1, 9V and 18C when comparing PCV20 with PCV13. These results highlight the importance of continuous monitoring of the distribution of serotypes in carriage and diseases as we enter the era of higher-valency PCVs. There would likely be a lag of a few years, as vaccinated children age into preschool and primary school, before any population-level impacts of reduced immunogenicity would be apparent. As more evidence based on various endpoints (IgG, memory B cells, colonization, and disease) from reduced-dose (1+1 and 0+1) schedules [28–30] and fractional dose regimens become available [31], our approach may also help answer similar questions about the relative effectiveness of various dosing regimens, with the prospect of considering duration of protection.

In addition to surveillance, future research efforts are indispensable in evaluating the impact of new vaccines. To confirm our estimates, the proposed model should be validated against real-world data as they accrue. To extend the comparisons included in our study to any pair of PCVs, network meta-analysis can help synthesize evidence. To translate these results to predict the impact on herd immunity, the duration of protection and transmission patterns should be considered, for instance, by using a dynamical transmission model. And primary data on immunogenicity and colonization should be collated from available clinical trials.

In conclusion, these analyses suggest that while new PCVs might provide sufficient protection against severe disease, protection against transmission might be somewhat reduced for some serotypes. This could have particular impacts on under-vaccinated age groups and other high-risk individuals. The overall impact and evolution with time should be evaluated in the local context, taking into account the proportion of fully vaccinated individuals in a given population. As such, further monitoring is critical to evaluate the impact of these changes in the coming years.

## Code & Data availability

Analysis code and data from the scoping review are available from: https://github.com/PneumococcalCapsules/PCV_colonization

## Ethics approval

The analyses of these de-identified data were exempted from review by the Yale University Institutional Review Boards (Protocol ID: 2000038285).

## Authors’ contribution

Conceived the study: DMW; designed and coded the models: AW, JLW and DMW; reviewed the literature: LF and SP; curated and cleaned the data: LF, SP and RD; analyzed the data: AW; supervised the analyses: DMW; contributed to methodology: AW, JLW, LF, SP, RD and DMW; interpreted the results: AW, JLW, LF, SP, RD and DMW; led project administration: DMW; provided resources: DMW; wrote the first draft of the manuscript: AW; revised the manuscript: AW, JLW, LF, SP, RD and DMW. All authors approved the final version of the submitted manuscript.

## Supporting information

eAppendices

## Data Availability

Analysis code and data from the scoping review are available from:

https://github.com/PneumococcalCapsules/PCV_colonization

## Acknowledgment

The research stay of AW was supported by the Max Planck Institute for Infection Biology. We thank Jessica Weaver, Josiah Thomas, and Maurice Ahsman for advice on the use of the conversion of ECL to ELISA unit based on their previous paper.

