## Supplementary material for "Estimating the serotype-specific association between the concentration of vaccine-induced serum antibodies and protection against pneumococcal colonization": eAppendices

| | |
|---|---|
| eAppendix 1 | Longitudinal data on post-PCV immunogenicity and colonization from Dagan et al. 2013 |
| eFigure 2 | Flowchart of the inclusion of clinical trials comparing PCV13 with PCV7 for the extraction of immunogenicity summary-level data |
| eFigure 3 | Flowchart of the inclusion of clinical trials comparing PCV15 with PCV13 for the extraction of immunogenicity summary-level data |
| eFigure 4 | Flowchart of the inclusion of clinical trials comparing PCV20 with PCV13 for the extraction of immunogenicity summary-level data |
| eAppendix 2 | Implementation of the change point model |
| eAppendix 3 | Sensitivity analysis of the change point model |
| eAppendix 4 | Details of the linear model |
| eAppendix 5 | Converting ECL-measured GMC to ELISA-measured GMC |
| eAppendix 6 | Pooling the estimated relative risk across trials using inverse-variance weighting |
| eFigure 15 | Absolute vaccine efficacy against colonization with 13 serotypes for PCV13, PCV15, and PCV20 calculated from the change point model's results |

**eAppendix 1. Longitudinal data on post-PCV immunogenicity and colonization from Dagan et al. 2013**

After excluding baseline colonization episodes and censoring colonization episodes with the same serotype following the first event, 2345 samples from 896 individuals (Bedouin 323; Jewish 573) in the longitudinal data on post-PCV immunogenicity and colonization [1,2] were included in the analysis. eTable 1 shows the serotype-specific measurements breakdown by serotype, ethnicity, and sex.

**eTable 1. A summary of data included in the analyses**

| | **Number of measurements** | **Number of colonizations** |
|---|---|---|
| **Total** | 28192 | 220 |
| **Serotype** | | |
| 4 | 2289 | 1 |
| 6B | 2064 | 16 |
| 9V | 2254 | 10 |
| 14 | 2208 | 19 |
| 18C | 2269 | 8 |
| 19F | 2061 | 37 |
| 23F | 2196 | 14 |
| 1 | 2279 | 2 |
| 3 | 2207 | 10 |
| 5 | 2162 | 0 |
| 6A | 2053 | 35 |
| 7F | 2149 | 2 |
| 19A | 2001 | 66 |
| **Ethnicity** | | |
| Bedouin | 10033 | 98 |
| Jewish | 18159 | 122 |
| **Sex** | | |
| Female | 14203 | 118 |
| Male | 13989 | 102 |

We excluded baseline colonization episodes, which were colonizations prior to the immunoglobulin G (IgG) measurement: those at 2, 4, and 6 months for post-booster IgG at age 7 months and those at 2, 4, 6, 7, and 12 months for post-booster IgG at age 13 months. eFigure 1 shows an example of observation inclusion for post-booster IgG at age 13 months. The individual was colonized by serotype 4 at baseline (12m); therefore, all of the observations for this serotype were excluded.

If a participant was found to be colonized by a given serotype, the subsequent observations for that same serotype were censored. In eFigure 1, the individual was colonized by serotype 6B at 18m, hence, the observations at 13m and 18m were included, and the observation at 24m was censored. The individual was not colonized by serotype 19F at baseline or at any follow-up visit; therefore, all observations (13, 18, and 24m) were included.

| Serotype | 12m | 13m | 18m | 24m |
|---|---|---|---|---|
| 4 | 1 | 1 | 0 | 0 |
| 6B | 0 | 0 | 1 | 0 |
| 19F | 0 | 0 | 0 | 0 |

1 = colonized
0 = not colonized

Censored observation

Eligible observation

**eFigure 1. A schematic for the inclusion of observations and the censoring of subsequent same-serotype colonization**

**eFigure 2. Flowchart of the inclusion of clinical trials comparing PCV13 with PCV7 for the extraction of immunogenicity summary-level data**

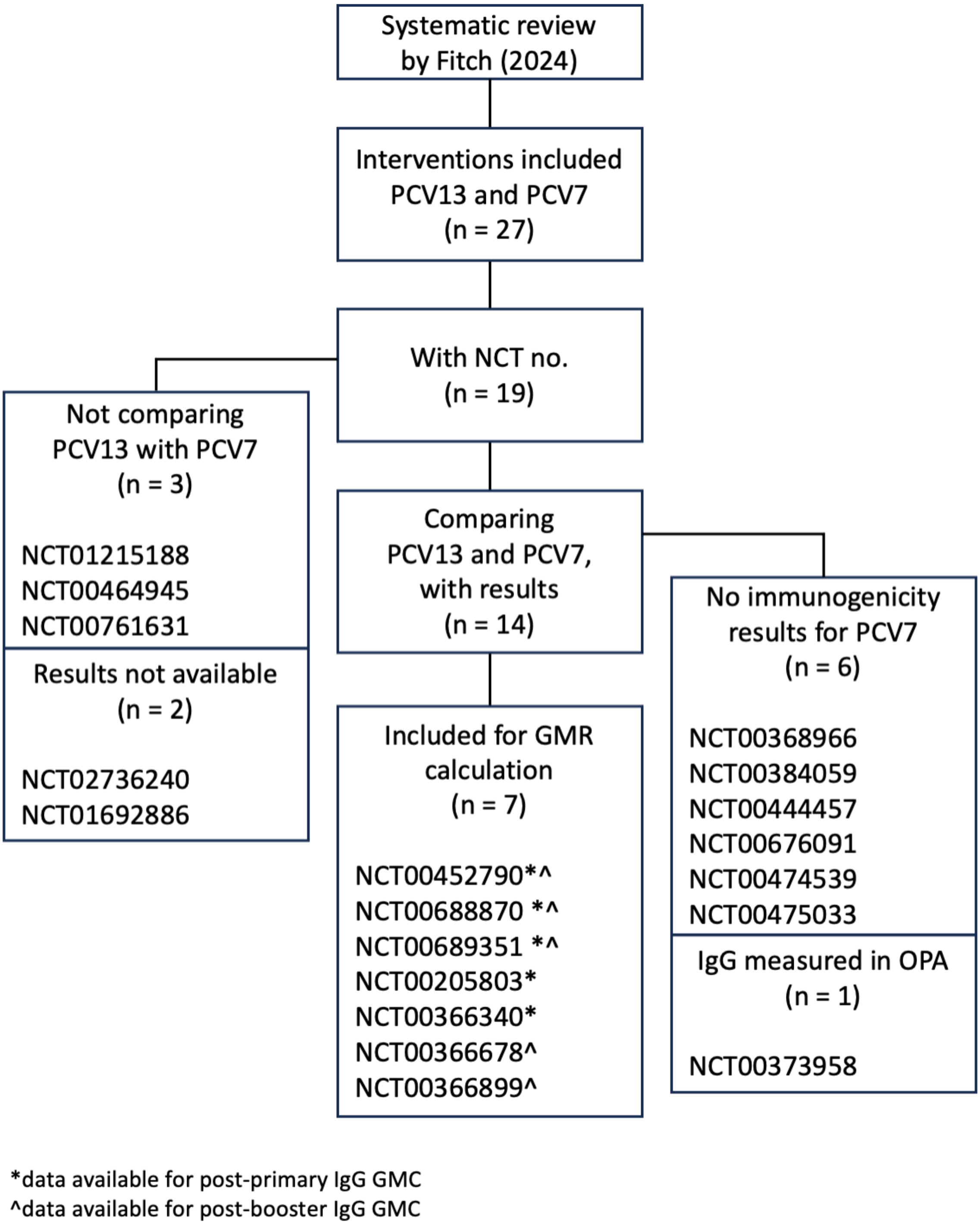

**eFigure 3. Flowchart of the inclusion of clinical trials comparing PCV15 with PCV13 for the extraction of immunogenicity summary-level data**

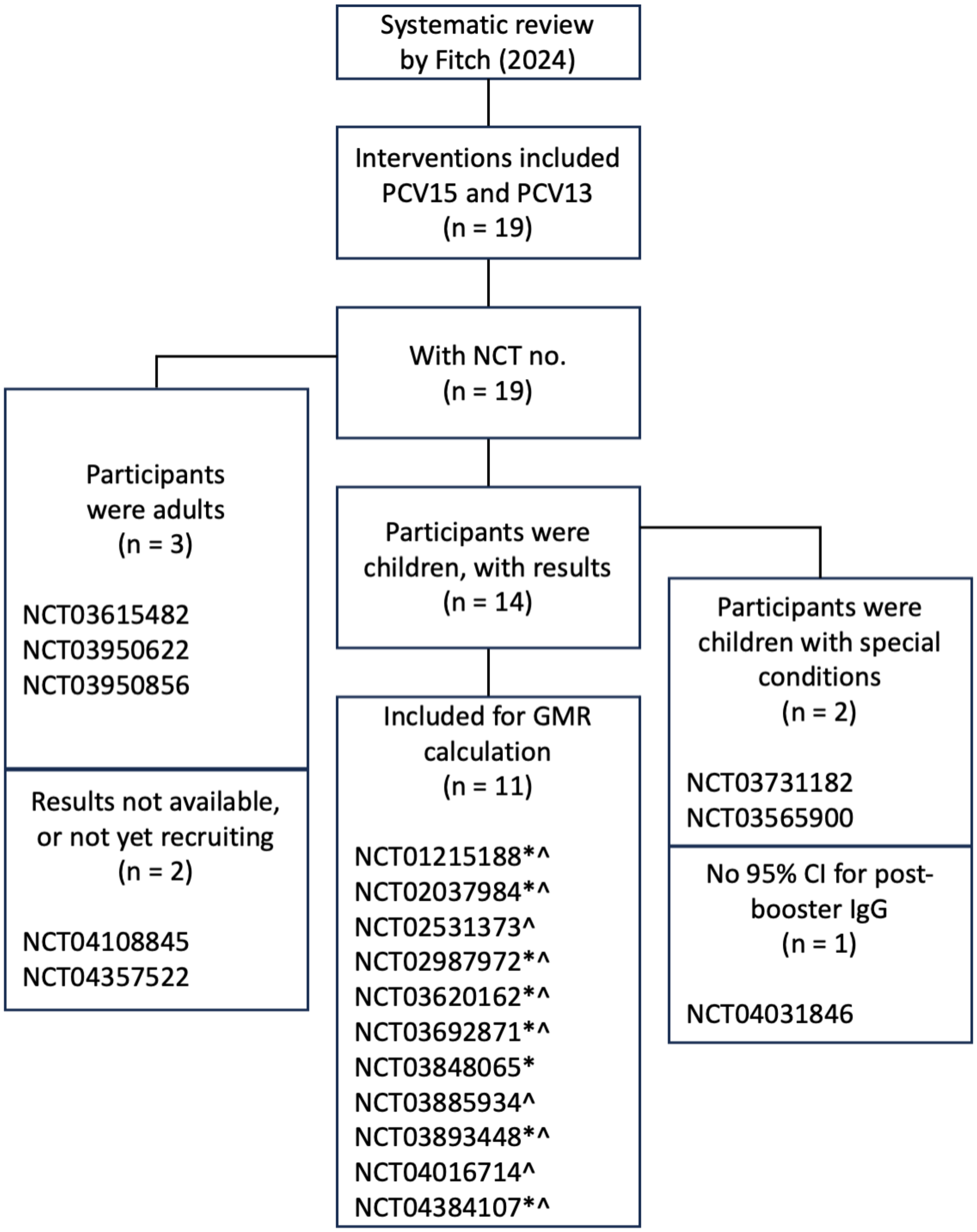


*data available for post-primary IgG GMC
^data available for post-booster IgG GMC

**eFigure 4. Flowchart of the inclusion of clinical trials comparing PCV20 with PCV13 for the extraction of immunogenicity summary-level data**

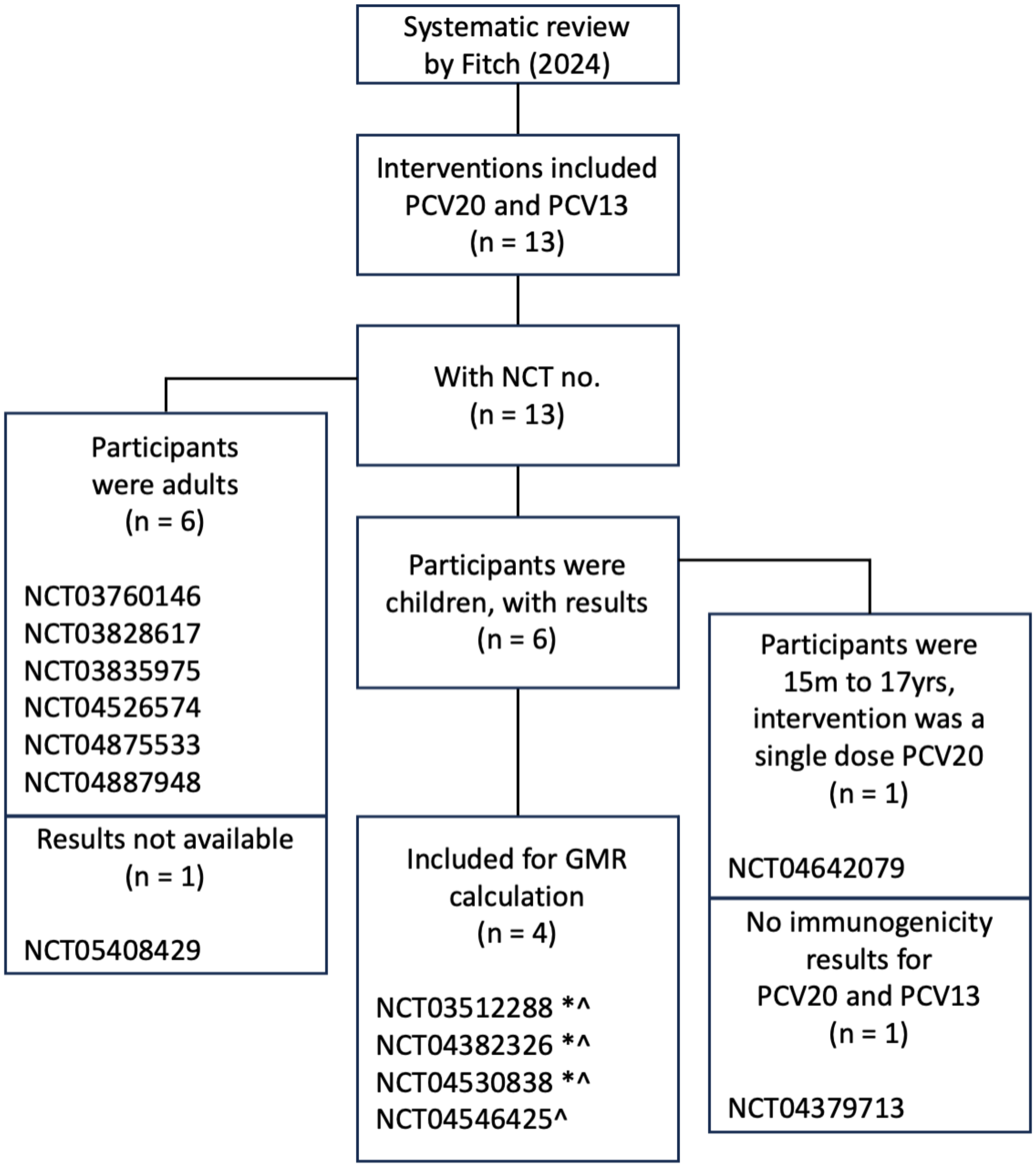


*data available for post-primary IgG GMC
^data available for post-booster IgG GMC

**eAppendix 2. Implementation of the change point model**

All analyses were performed in RStudio with R version 4.2.2 (R) [3] using the package "rjags" version 4–14 [4]. The change point model was implemented with three independent chains for 10000 iterations each after discarding the first 10000 samples of each chain prior to convergence of the model, resulting in 30000 samples in total. Model convergence was assessed by visual inspection of trace plots, with no obvious signs of problems being observed. The posteriors of change points for serotypes 4, 9V, 18C, 1, 5, and 7F showed larger variability, potentially due to the low event counts for these serotypes (eTable 1). The variability in the estimates for each serotype, stratified by ethnicity, is visualized in eFigure 5; the model curves scaled to 0 to 1 by the maximum risk for each serotype is visualized in eFigure 6 for easier comparison of change points across serotypes.

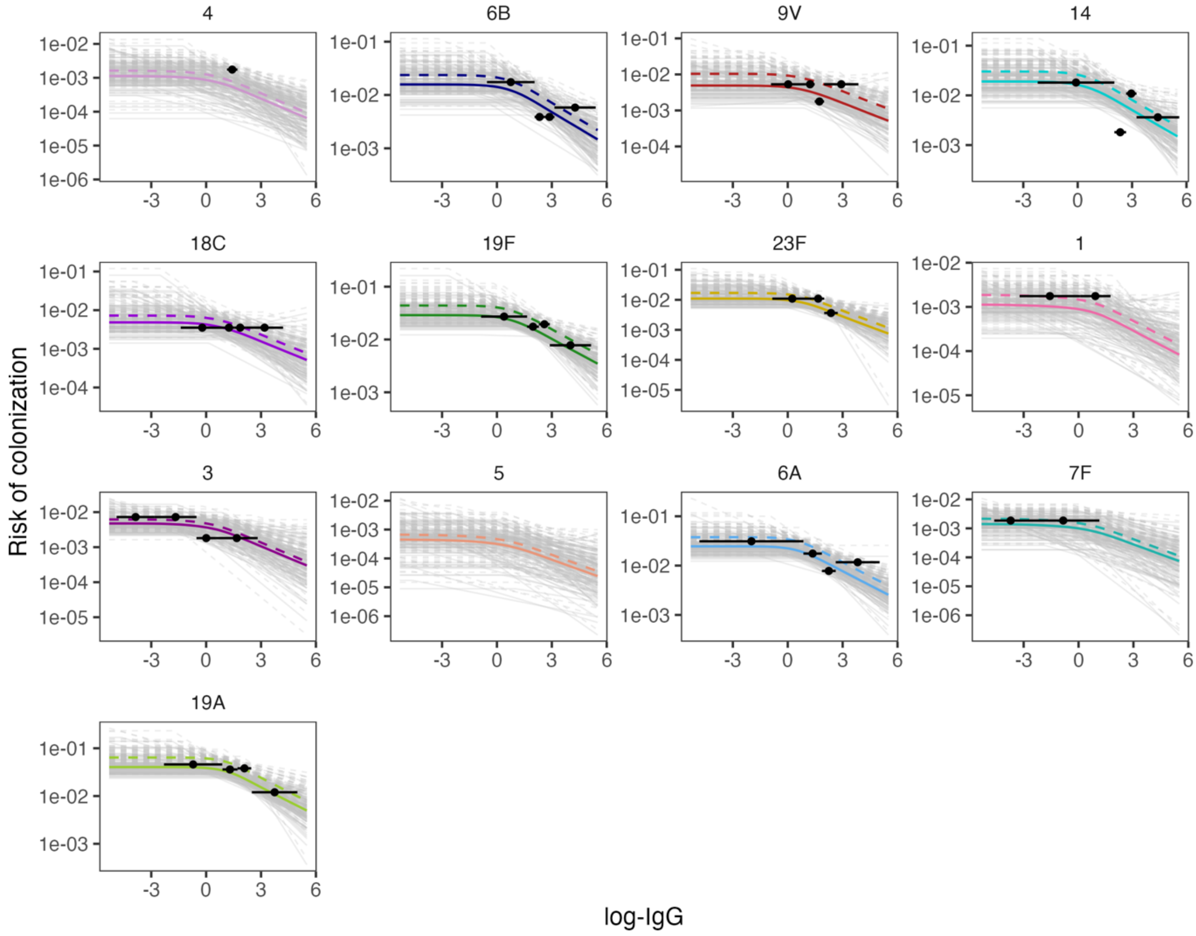


**eFigure 5. The observed proportion of colonization and predicted risk of colonization by antibody concentration for each serotype**

In each panel, the colored lines indicate the median predicted risk of colonization; the grey lines represent 100 estimates randomly drawn from the posterior distribution, visualizing the uncertainty of the estimates. Solid lines represent the estimates for the Jewish population; dashed line represent the estimates for the Bedouin population. For each serotype, we cut the observed immunoglobulin G (IgG) concentration into four quartiles, and plotted the observed proportion of colonization against the average IgG concentration in each quartile using a black point, with the error bars indicating the range of the IgG concentration in a quartile.

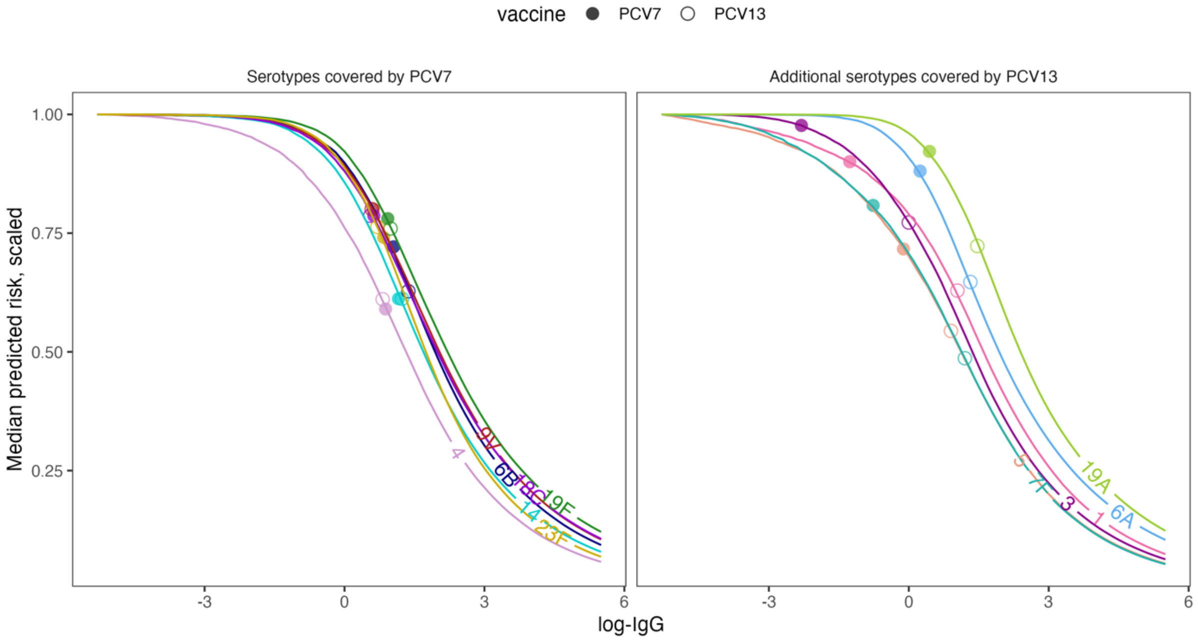


**eFigure 6. The predicted probability of colonization by antibody concentration scaled to 0 to 1 by the maximum risk for each serotype**

The median predicted risk of colonization acquisition from the hierarchical Bayesian model, scaled to 0 to 1 by the maximum risk for each serotype (left: serotypes covered by PCV7 – 4, 6B, 9V, 14, 18C, 19F, 23F; right: additional serotypes covered by PCV13 – 1, 3, 5, 6A, 7F, 19A), is plotted against the concentration of serum antibodies. The points indicate the antibody concentrations achieved by the booster dose of PCV7 (filled circle) and PCV13 (open circle), as an inverse-variance weighted average of the immunoglobulin G (IgG) geometric mean concentration (GMC) reported by the head-to-head clinical trials.

### eAppendix 3. Sensitivity analysis of the change point model

First, we assumed that if there is a change point for a given serotype, the antibody concentration (log-IgG) corresponding to the change point would fall in the observed range of log-IgG. Therefore, we used the mean and variance of the observed log-IgG as the prior and assumed $\mu_{cp}$ to be normally distributed with mean equal to the log-IgG and variance equal to the variance of log-IgG in the main analysis.

To test a less informative prior, we assumed $\mu_{cp}$ to be normally distributed with mean 0 with variance $10^4$ in this sensitivity analysis. Although the modeled risk curves show slightly different shapes compared to the model in the main analysis (eFigure 7), the uncertainty remained similar (eFigure 8). In general, the change points appeared to be slightly more subtle when using a less informative prior. This has no impact on the relative risk inference for all serotypes across the vaccine comparisons, except for serotype 3 when comparing PCV15 vs. PCV13 and PCV20 vs. PCV13 (eFigure 9). The main analysis did not find a difference in protection against serotype 3 colonization comparing PCV15 and PCV20 with PCV13. When using a less informative prior, the protection against colonization with serotype 3 was estimated to be better for PCV15 than PCV13 and worse for PCV20 than PCV13.

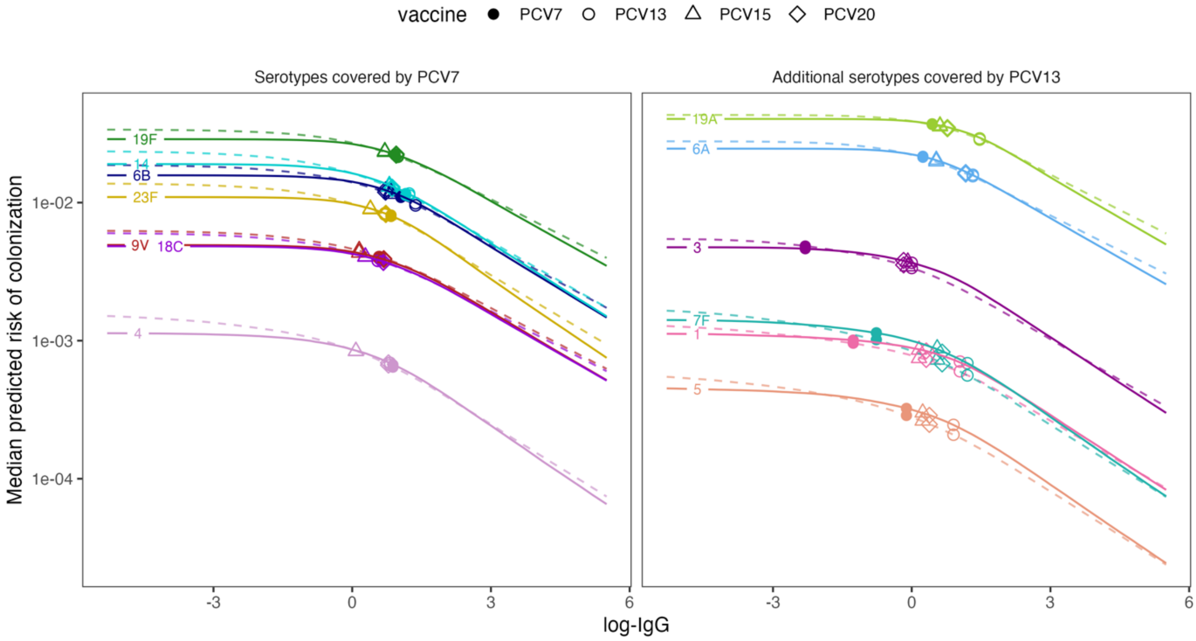


**eFigure 7. A comparison of the predicted probability of colonization by antibody concentration for each serotype using informative prior vs. alternative prior for $\mu_{cp}$**

The median predicted risk of colonization acquisition from the hierarchical Bayesian model for each serotype (left: serotypes covered by PCV7 – 4, 6B, 9V, 14, 18C, 19F, 23F; right: additional serotypes covered by PCV13 – 1, 3, 5, 6A, 7F, 19A) is plotted against the concentration of serum antibodies. Solid lines represent the model in the main analysis, which used an informative prior for $\mu_{cp}$; dashed lines represent the model in this sensitivity analysis, which used a weakly informative prior for $\mu_{cp}$. The points indicate the antibody concentrations achieved by the booster dose of PCV7 (filled circle), PCV13 (open circle), PCV15 (open triangle), and PCV20 (open diamond), as an inverse-variance weighted average of the immunoglobulin G (IgG) geometric mean concentration (GMC) reported by the head-to-head clinical trials. The y-axis is displayed in the log10 scale.

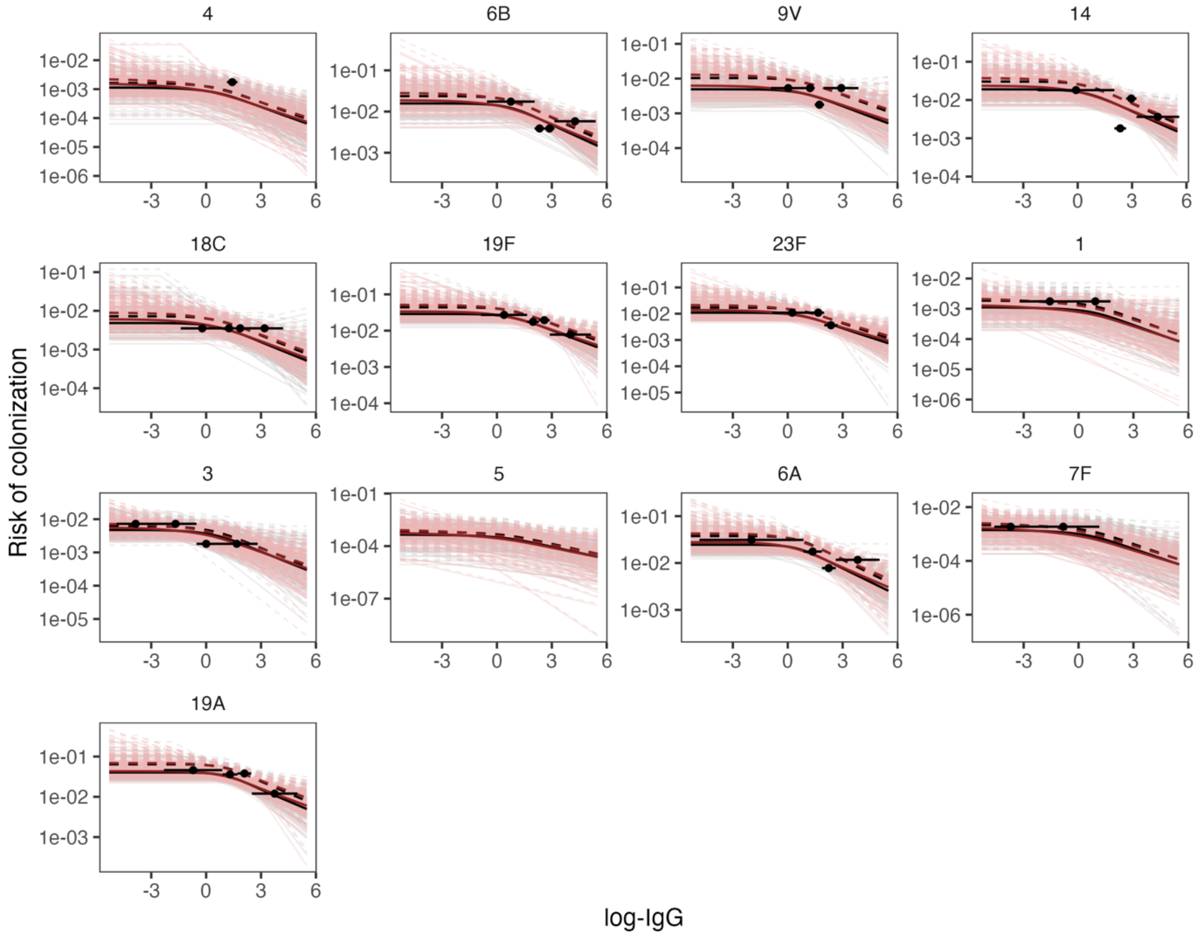


**eFigure 8. The observed proportion of colonization and predicted risk of colonization by antibody concentration for each serotype from the change point models using informative prior vs. alternative prior for $\mu_{cp}$**

In each panel, the bold lines indicate the median predicted risk of colonization and the lines in the background represent 100 estimates randomly drawn from the posterior distribution, visualizing the uncertainty of the estimates. Solid lines represent the estimates for the Jewish population; dashed line represent the estimates for the Bedouin population. The set of lines with a black/grey theme color came from the model in the main analysis, which used an informative prior for $\mu_{cp}$, and the ones with a dark red/pink theme color came from the model in this sensitivity analysis, which used a weakly informative prior for $\mu_{cp}$. For each serotype, we cut the observed immunoglobulin G (IgG) concentration into four quartiles, and plotted the observed proportion of colonization against the average IgG concentration in each quartile using a black point, with the error bars indicating the range of the IgG concentration in a quartile.

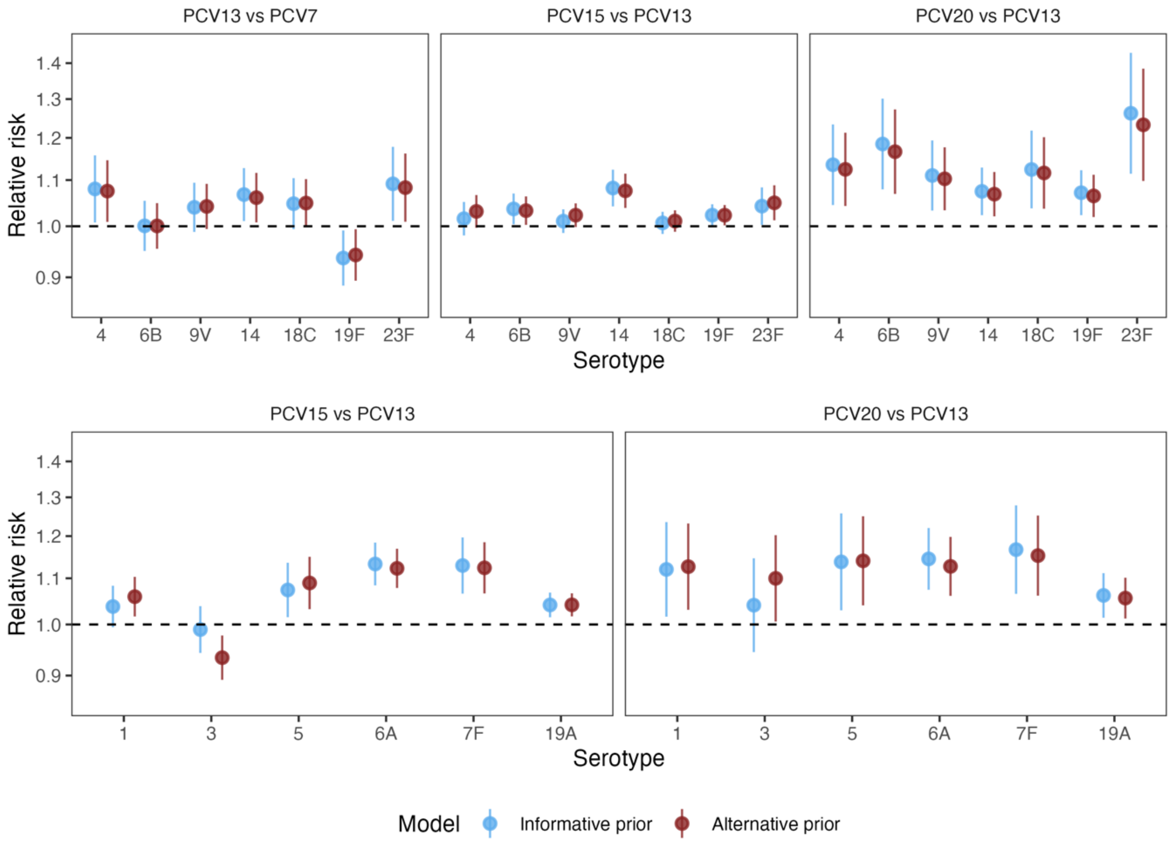


**eFigure 9. Relative risk of colonization with 13 serotypes predicted by the change point models using informative prior vs. alternative prior for $\mu_{cp}$ (PCV13vs7, PCV15vs13, PCV20vs13)**

**eAppendix 4. Details of the linear model**

Before using the change point model, we initially built a hierarchical Bayesian linear model that assumed a log-log linear relationship between the risk of colonization and serum concentration of immunoglobulin G (IgG) using the R package "INLA" version 23.04.24 [5]. The relative risk of colonization after being vaccinated with a higher-valency pneumococcal conjugate vaccine (PCV2) to a lower-valency one (PCV1) can be estimated using the formula:

$$\ln(\mathrm{RR}) = \beta_1 \ln(\mathrm{GMR})$$

where

- $\beta_1$ is the serotype-specific coefficient; and
- ln(GMR) is the natural-logarithm of the GMC-ratio (GMR) of PCV2 : PCV1, this ratio is given by log(GMC_PCV2 / GMC_PCV1) where GMC_PCV2 is the antibody level following immunization with PCV2 and GMC_PCV1 is the antibody level following immunization with PCV1.

The estimated relative risk of colonization with the 13 serotypes covered by PCV13 from the linear model is shown in eFigure 10, stratified by vaccine dose (post-primary and post-booster), ethnicity and vaccine comparison. The estimates of the relationship between IgG concentration and risk of colonization for the Bedouin and Jewish groups were not notably different; therefore, we only accounted for the baseline colonization risk difference by including the ethnicity-specific intercepts in the change point model. eFigure 11 shows a comparison of the estimates from the linear model and the change point model.

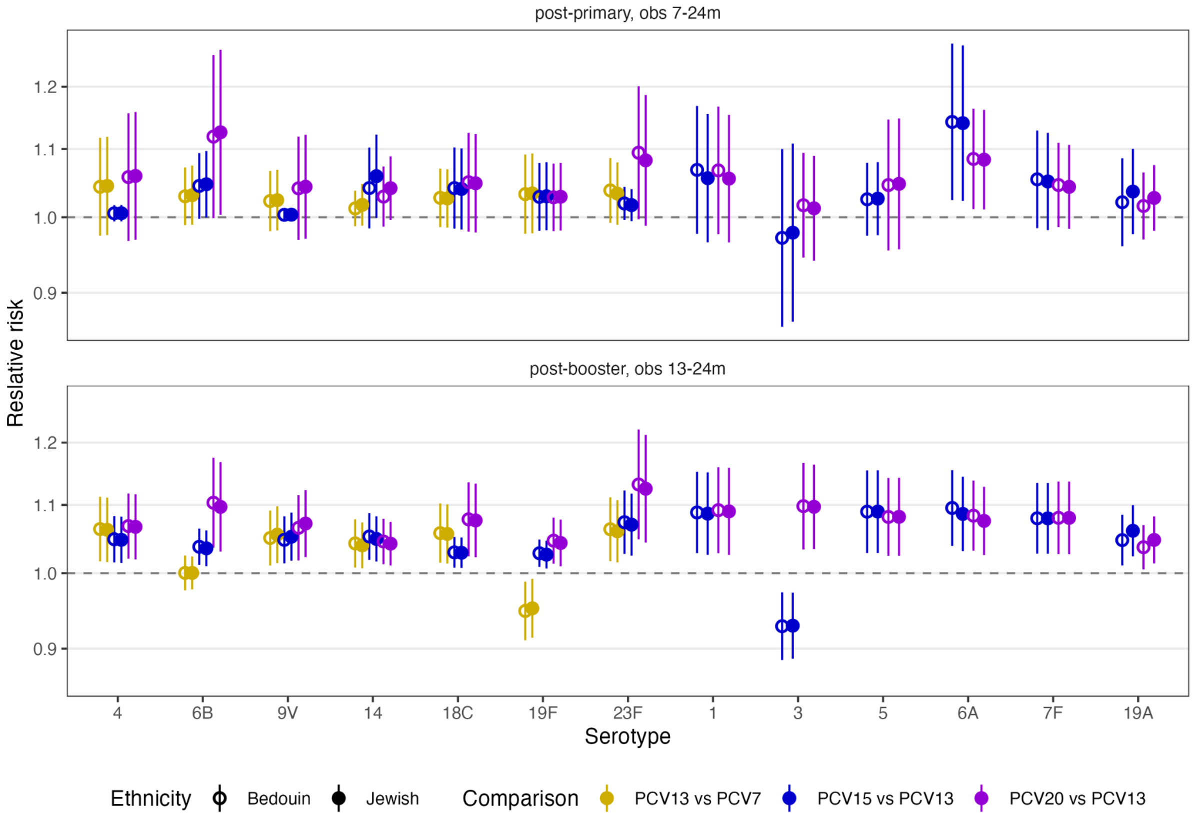


**eFigure 10. Relative risk of colonization with 13 serotypes predicted by the linear model, stratified by ethnicity (PCV13vs7, PCV15vs13, PCV20vs13)**

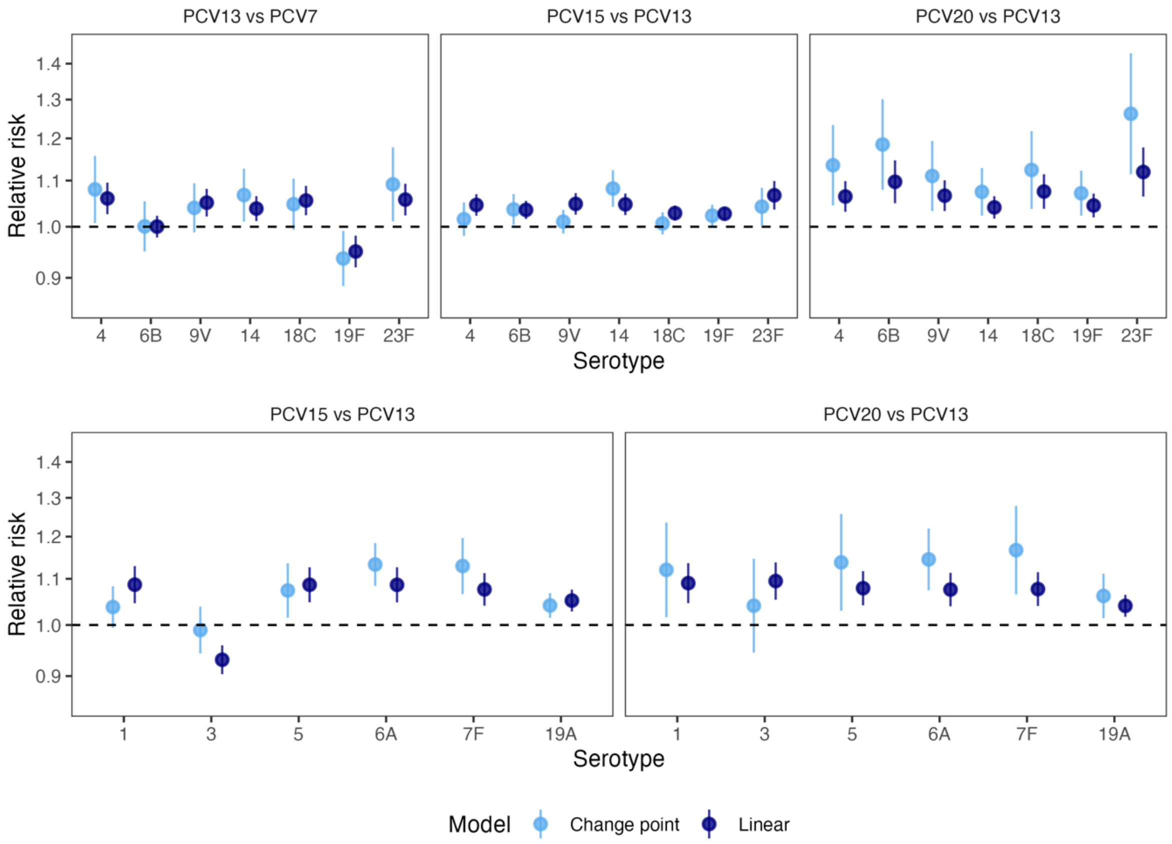


**eFigure 11. Relative risk of colonization with 13 serotypes predicted by the linear model and the change point model (PCV13vs7, PCV15vs13, PCV20vs13)**

**eAppendix 5. Converting ECL-measured GMC to ELISA-measured GMC**

To convert the geometric mean concentration (GMC) measured by an electrochemiluminescence (ECL) assay to GMC measured by enzyme-linked immunosorbent assay (ELISA), we use the formula [6]:

$$ELISA_i = \left(\frac{ECL}{e^{\alpha_i}}\right)^{\frac{1}{\beta_i}}$$

where $\alpha_i$ and $\beta_i$ represents the intercept and slope for the relationship between the ECL assay and ELISA for each serotype *i*.

The slopes $\beta_i$ were obtained directly from Table 2 in [7], whereas the intercepts can be calculated by:

$\alpha_i$ = log(fold difference * 0.35/ (0.35 ^ ($\beta_i$ / 1)))

**eAppendix 6. Pooling the estimated relative risk across trials using inverse-variance weighting**

For each serotype, we estimated the relative risk (RR) of colonization by combining the reported post-booster immunoglobulin G (IgG) geometric mean concentration (GMC) with 95% confidence intervals from each head-to-head trial and the change point model. The point estimate $RR_i$ is plotted with a black point in eFigures 12–14, with the 95% credible intervals indicated by error bars.

We then assigned a weight $W_i$ to the estimate $RR_i$ of each study $i$:

$$W_i = \frac{1/\sigma_i^2}{\sum_{i=1}^{n} 1/\sigma_i^2}$$

where $1/\sigma_i^2$ is the inverse-variance of the estimate $RR_i$, and calculated the weighted average $\widehat{RR}$ for each serotype:

$$\widehat{RR} = \sum_{i=1}^{n} RR_i \times W_i$$

The pooled estimate $\widehat{RR}$ is plotted with an open diamond in eFigures 12–14.

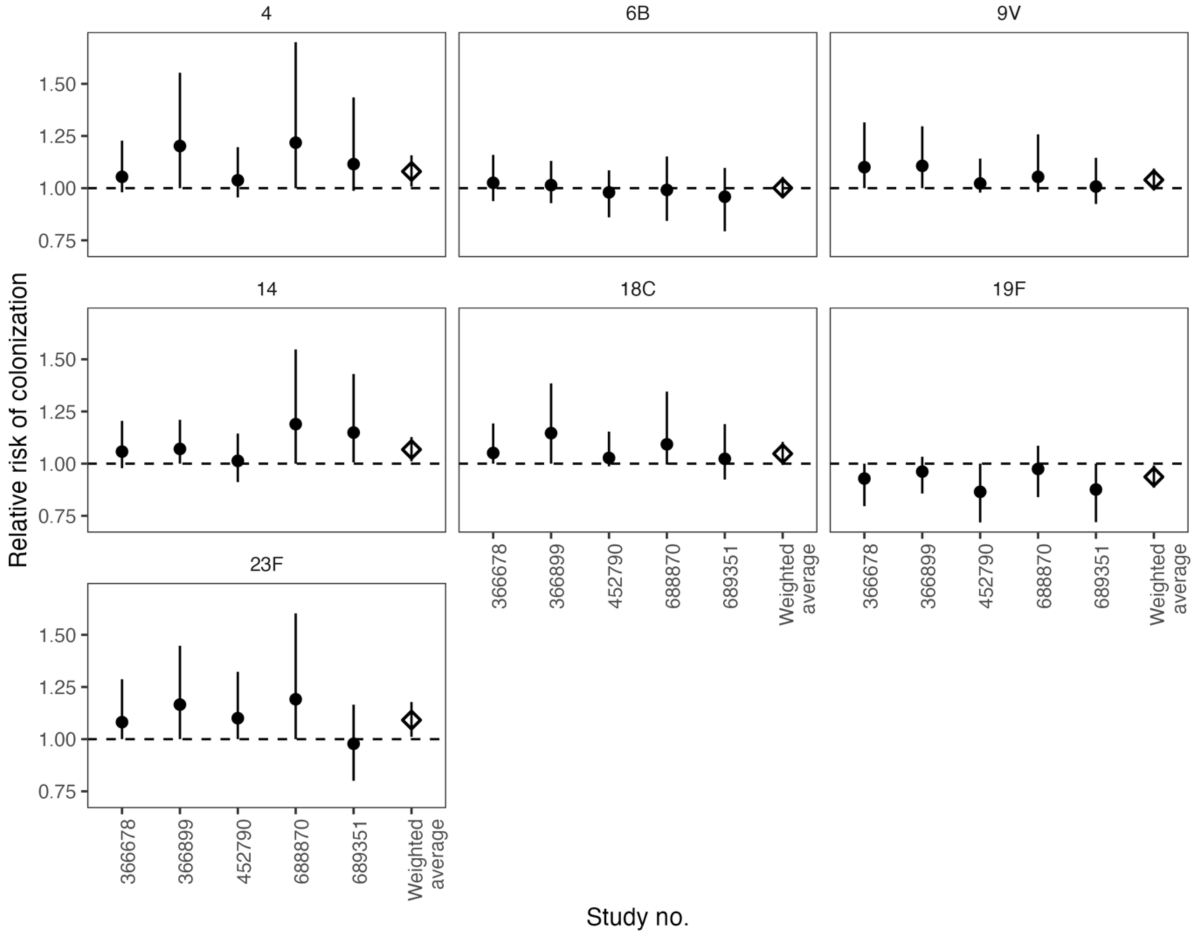


**eFigure 12. Relative risk of colonization with 7 serotypes predicted by the change point model in each study and the inverse-variance weighted average (PCV13vs7)**

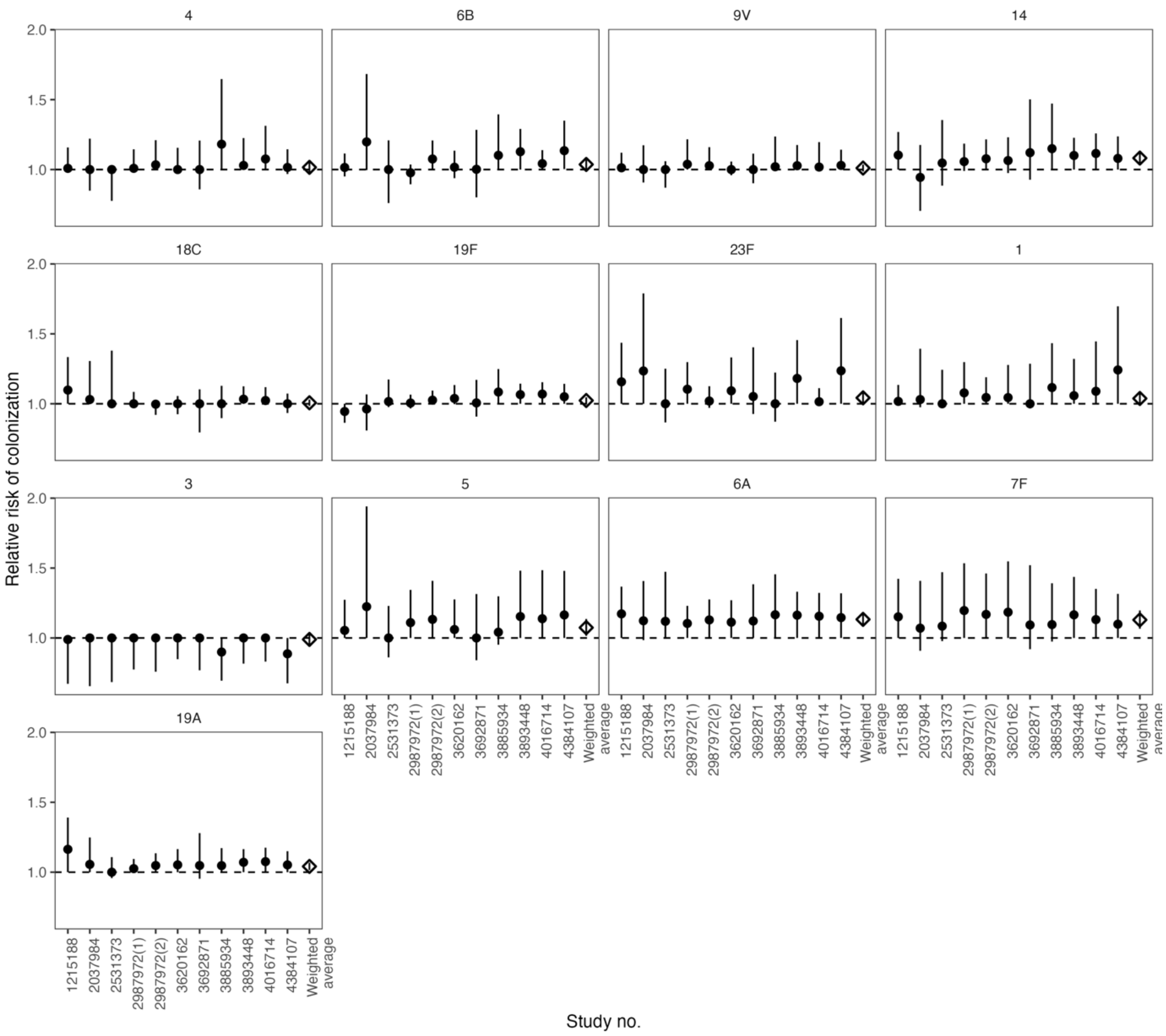


**eFigure 13. Relative risk of colonization with 13 serotypes predicted by the change point model in each study and the inverse-variance weighted average (PCV15vs13)**

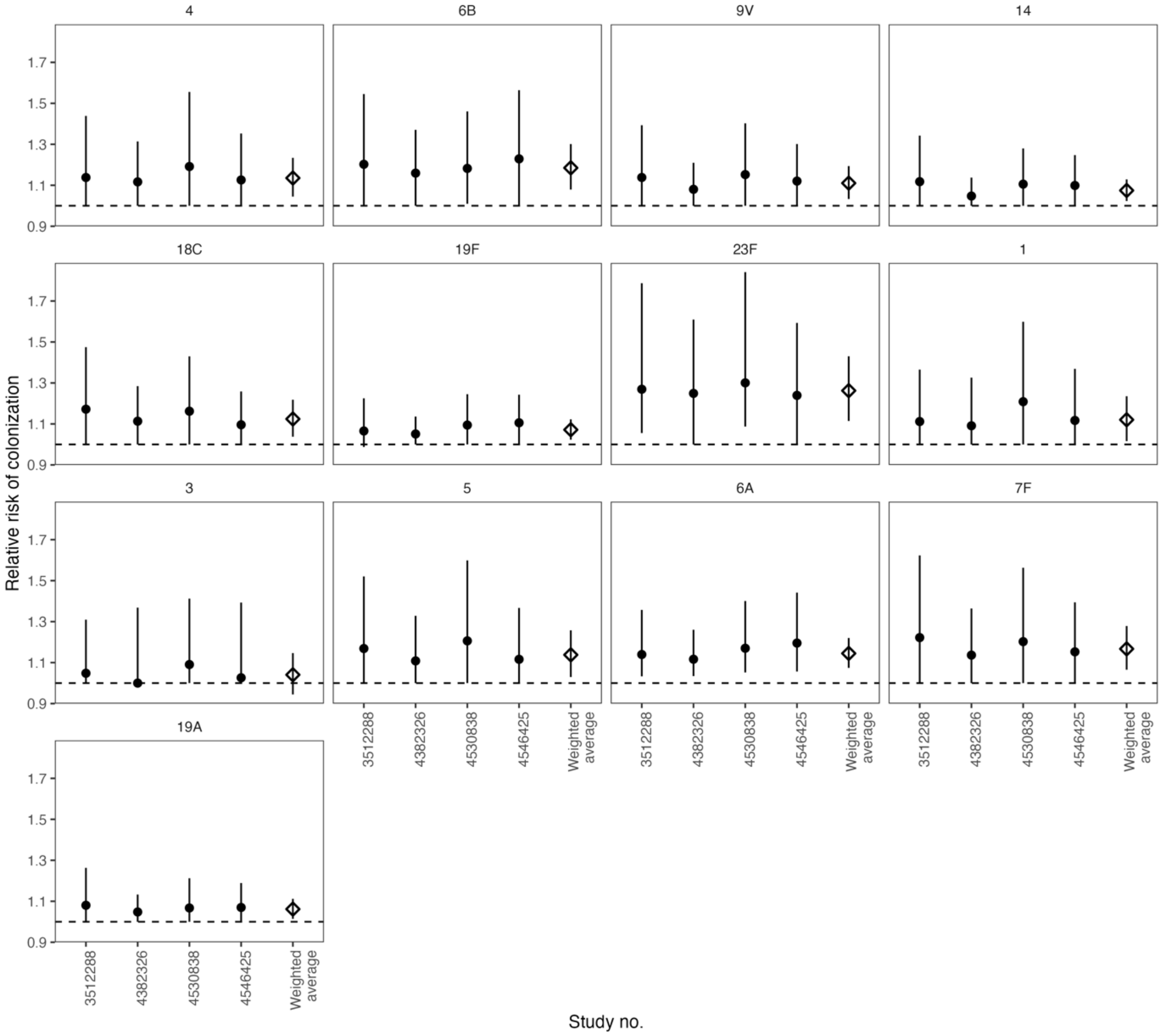


**eFigure 14. Relative risk of colonization with 13 serotypes predicted by the change point model in each study and the inverse-variance weighted average (PCV20vs13)**

**eFigure15. Absolute vaccine efficacy against colonization with 13 serotypes for PCV13, PCV15, and PCV20 calculated from the change point model's results, assuming the vaccine effectiveness of PCV7 to be 60% (an arbitrary reference number based on [8,9])**

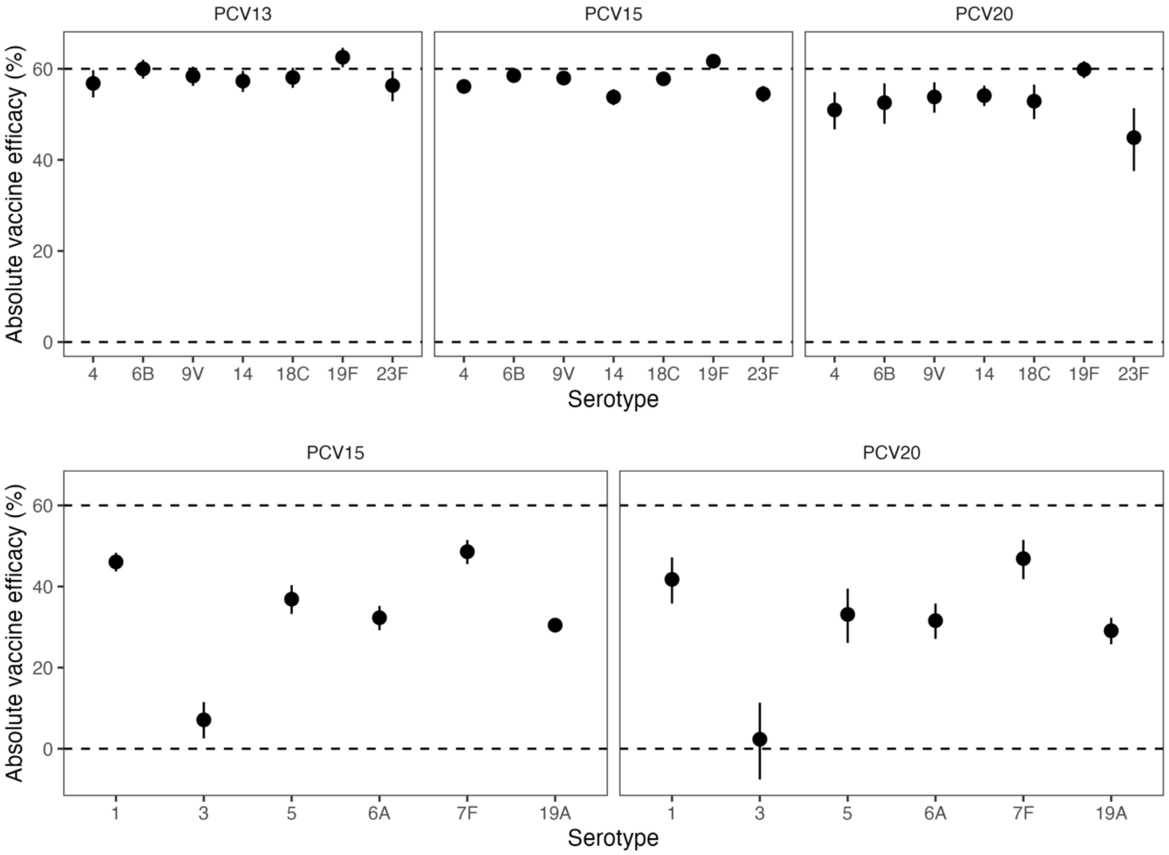
